# Posttraumatic stress disorder is associated with alpha dysrhythmia across the visual cortex and the default mode network

**DOI:** 10.1101/19011841

**Authors:** Kevin J. Clancy, Jeremy A. Andrzejewski, Mingzhou Ding, Norman B. Schmidt, Wen Li

## Abstract

**Background:** Anomalies in default mode network (DMN) activity and alpha (8-12 Hz) oscillations have been independently observed in posttraumatic stress disorder (PTSD). Recent spatiotemporal analyses suggest that alpha oscillations support DMN functioning via inter-regional synchronization and sensory cortical inhibition. Therefore, we examined a unifying pathology of alpha deficits in the visual-cortex-DMN system in PTSD.

**Methods:** Patients with PTSD (*N* = 25) and two control groups—patients with Generalized Anxiety Disorder (*N* = 24) and healthy controls (*N* = 20)—underwent a standard eyes-open resting state (S-RS) and a modified resting state (M-RS) of passively viewing salient images (known to deactivate the DMN). High-density electroencephalogram (hdEEG) were recorded, from which intracortical alpha activity (power and connectivity/Granger causality) was extracted using the exact low-resolution electromagnetic tomography (eLORETA).

**Results:** Patients with PTSD (vs. controls) demonstrated attenuated alpha power in the visual cortex and key hubs of the DMN (posterior cingulate cortex/PCC and medial prefrontal cortex/mPFC) at both states, the severity of which further correlated with hypervigilance symptoms. With increased visual input (at M-RS vs. S-RS), patients with PTSD further demonstrated reduced alpha-frequency directed connectivity within the DMN (PCC→mPFC) and, importantly, from the visual cortex (VC) to both DMN hubs (VC→PCC and VC→mPFC), linking alpha deficits in the two systems.

**Conclusions:** These interrelated alpha deficits align with DMN hypoactivity/hypoconnectivity, sensory disinhibition, and hypervigilance in PTSD, representing a unifying neural underpinning of these anomalies. The identification of visual-cortex-DMN alpha dysrhythmia in PTSD further presents a novel therapeutic target, promoting network-based intervention of neural oscillations.

## INTRODUCTION

Posttraumatic stress disorder (PTSD) is a highly debilitating psychiatric disorder. Prevailing models of PTSD neuropathology have focused on dysfunctions of the prefrontal-cortex-amygdala circuit (1, 2). Recently, evidence has further extended this circuit pathology to implicate large-scale brain network anomalies in the neuropathology of PTSD (1, 3-6). Anomalies in the default mode network (DMN), a major resting-state network (RSN), have been especially highlighted in this literature, characterized by attenuated network activity and disrupted network communication, involving reduced within-network and increased cross-network connectivity (4, 7-15).

The DMN is one of the most consistently identified RSNs, anchored in a midline core consisting of two key hub structures—the posterior cingulate cortex (PCC) and the medial prefrontal cortex (mPFC) (16-18). While prominent in the resting state, the DMN is deactivated by salient sensory input or externally-oriented cognitive processing and, accordingly, exhibits reciprocal inhibition with networks associated with those processes (e.g., the visual, salience, and dorsal attention networks) (19-22). As such, DMN dysfunctions can, on one hand, interrupt internal mentation and ‘tranquil’ resting states and, on the other, heighten visual vigilance and attention. Indeed, DMN dysfunction has been linked to heightened emotion/stress response and weakened top-down regulation, perpetuating stress symptoms (18, 23, 24) and potentially underpinning PTSD symptoms of hypervigilance and negative intrusions (7, 25, 26).

While neuroimaging data have isolated the DMN as the dominant network in the resting brain, electrophysiological data (electroencephalogram/EEG or magnetoencephalogram/MEG) have identified alpha (8-12 Hz) oscillations as the dominant electrical activity in the resting brain (27-29). Alpha oscillations represent a key neural mechanism in mediating long-range inter-regional interactions (27, 30, 31). Importantly, converging evidence has linked alpha oscillations directly to DMN activity (especially in its hubs of PCC and mPFC), advancing the notion that the DMN could be organized and maintained by long-range synchronization of alpha oscillations (32-39).

While alpha oscillations positively correlate with DMN activity, they are known to negatively correlate with visual network activity (27, 28, 39-41), consistent with the inverse relationship between the two networks. Alpha oscillations (originating in the sensory cortex and thalamus) play a key role in visual inhibition by suppressing visual cortical excitation and feedforward propagation (27-31, 40, 42-47). This active function of alpha oscillations can present a second mechanism—visual cortical inhibition—to support DMN activity. That is, by suppressing visual processing and propagation, alpha oscillations could protect the DMN from environmental disruptions, especially in busy sensory environments.

Alternatively, deficient alpha activity could give rise to DMN dysfunctions by failing to sustain long-range synchronization across the DMN and failing to inhibit sensory afferents to the DMN. Aberrant alpha oscillations have been featured in a “thalamocortical dysrhythmia” model of neuropsychiatric disorders (48, 49), which are conceptualized transdiagnostically as oscillopathies (50, 51). Recently, our lab demonstrated reduced resting-state alpha activity (in both power and long-range connectivity) in patients with PTSD (vs. healthy controls and patients with generalized anxiety disorder/GAD), which was further associated with severity of hypervigilance (52). Given the demonstrated association between alpha oscillations and DMN activity, we hypothesized that PTSD is associated with deficient alpha activity in the DMN and deficient alpha-mediated inhibition of sensory cortical input to the DMN.

Therefore, extracting intracortical alpha activity using the exact low-resolution electromagnetic tomography (eLORETA) (53) of high-density EEG, we conducted source-level analysis of resting-state alpha activity in patients with PTSD, relative to healthy controls and patients with GAD. The GAD group was included to rule out effects of general anxiety and hyperarousal that would confound resting-state neural oscillations (54). To highlight the vulnerability of the DMN to salient sensory input, we included a modified resting state (M-RS) involving passive viewing of salient pictures, in addition to a standard, eyes-open resting state (S-RS). We assessed two specific hypotheses of alpha deficits in PTSD, including (1) a deficit in alpha activity (i.e., attenuated alpha power and connectivity) in the DMN and (2) a deficit of alpha inhibition of visual cortical (VC) activity (i.e., attenuated alpha power) and relay to the DMN (i.e., attenuated alpha-frequency directed connectivity; VC→DMN), especially with strong visual input (as in the M-RS). Finally, we examined clinical associations between such deficits and the PTSD symptom of hypervigilance, linking neuropathology to clinical symptomatology.

## METHODS & MATERIALS

### Participants

The data presented here belong to a larger study, part of which was initially reported in Clancy et al. 2017 (52). Participants consisted of outpatients with a diagnosis of PTSD (*n* = 25) or GAD (*n* = 24), and healthy controls (HC; *n* = 20). Participants were matched for age and gender across groups. All participants had no neurological disorders or history of severe traumatic brain injury. All participants provided written, informed consent to participate in the study, which was approved by both the Florida State University Institutional Review Board and the Department of Defense Human Research Protection Official’s Review. Demographic details are presented in Table 1, and in greater detail in Clancy et al. 2017 (52).

**Table 1.** Participant Demographics.

|  | PTSD | Controls |
| --- | --- | --- |
| Age (years) | 34.6 ± 10.4 | 31.1 ± 13.1 |
| Gender (female/male) | 16/9 | 25/19 |
| Substance Use (%) <sup>†</sup> | 64%* | 7.5% |
| Medication Use (%) | 40% | 36% |
| PCL Total | 61.0 ± 16.1* | 37.6 ± 13.4 |
| PCL-Hypervigilance | 4.1 ± 1.0* | 2.1 ± 1.2 |
| BAI | 26.2 ± 15.7* | 11.8 ± 9.4 |
| BDI | 26.8 ± 12.5* | 17.4 ± 9.7 |

PCL = Posttraumatic Stress Disorder Checklist, BAI = Beck Anxiety Inventory, BDI = Beck
Depression Inventory; <sup>†</sup> = Subjects with opioid, stimulant, and cocaine use were excluded. \* $p < 0.005$ .

### Clinical Assessment

Diagnoses were assessed by trained clinicians using the Structured Clinical Interview for DSM-V (55). Participants additionally completed the PTSD Checklist for DSM-IV: civilian version (56). In the present study, internal consistency was high for the total questionnaire (α = 0.95) and the hyperarousal subscale (α = 0.84). As in previous work (52, 57), Item 16 assessing symptoms of hypervigilance or ‘being watchful or on guard’ was extracted to index hypervigilance.

### Experimental Paradigm

Participants were seated in a comfortable recliner in a dimly lit, sound attenuated and electrically shielded room. High-density electroencephalogram (EEG) data were recorded during two resting states. To evaluate intrinsic neural activity, a standard resting state (S-RS) recording was conducted first, lasting 2 minutes while participants fixated on a crosshair on the screen. To assess the impact of environmental sensory input on alpha oscillations and DMN functioning, a modified resting state (M-RS) recording was also conducted, involving 5 minutes of passively viewing a continuous stream of images (subtending a visual area of 7.8° X 5.8°), each for 333 ms. Images were chosen from the International Affective Picture System (58), depicting neutral (e.g. buildings, daily objects; n = 322), positive (e.g. erotic; n = 253), and negative (e.g. mutilation; n =346) scenes, randomly intermixed.

### EEG Acquisition and Preprocessing

EEG data were recorded from a 96 channel BrainProducts actiCap system with Neuroscan SynAmps RT amplifiers (1000 Hz sampling rate, 0.05-200 Hz online bandpass filter, referenced to the FCz channel). Electro-oculogram (EOG) was recorded using four electrodes with vertical and horizontal bipolar derivations. EEG/EOG data were downsampled to 250 Hz, high-pass (1 Hz) and notch (60 Hz) filtered. We then applied *Fully Automated Statistical Thresholding for EEG artifact Rejection* algorithm (FASTER) (59) for artifact detection, correction, and rejection. Output data were epoched into 1-second segments and submitted to eLORETA for source analyses. Details regarding the FASTER procedure are outlined in the Supplemental Material.

### Exact low-resolution electromagnetic tomography (eLORETA)

Using the high-density, artifact-minimized EEG data, we conducted intracranial source analyses using eLORETA, a linear inverse solution to reconstruct cortical activity with scalp EEG data (53). The LORETA algorithm has been cross-validated in multiple studies combining EEG-based LORETA with fMRI (60-64), positron emission tomography (65, 66), and intracranial recordings (67). The solution space consists of 6239 cortical gray matter voxels with a spatial resolution of 5 x 5 x 5 mm in a realistic head model. eLORETA is a suitable tool to investigate network activity and connectivity (36, 68-70) and has provided important network insights into psychiatric disorders (36, 54, 71).

For accurate inverse solutions, eLORETA was modeled for the S-RS and M-RS separately, from which whole-brain source estimates of alpha (8-12 Hz) power were derived (53). For source-based alpha-frequency connectivity analysis, time series of regions of interest were derived from eLORETA (36), which were then submitted to Granger causality analysis based on bivariate autoregressive (AR) modelling (72). A model order of 20 (80 ms in time for a sampling rate of 250 Hz) was chosen in a two-step process: (1) Akaike Information Criterion (AIC) and (2) comparing spectral estimates obtained by the Fourier-based AR model for data pooled across all subjects (44).

Regions of Interest (ROIs): DMN hubs (i.e., PCC and mPFC) and visual cortex were selected as ROIs (16, 73). ROIs were defined by grey-matter voxels within a 10 mm radius of the ROI centroids (53, 71). All ROIs were centered on the midline to incorporate both hemispheres, with centroid coordinates obtained from the Neurosynth (www.neurosynth.org) meta-analysis maps (as peak voxels) of “default mode” (for the PCC: 0, −50, 30 and the mPFC: 0, 50, 0) and “passive viewing” (for the visual cortex: 0, −90, 20). All coordinates are reported in Montreal Neurological Institute (MNI) space. For connectivity analysis, both directions for each pair were examined such that the three ROIs resulted in a 3 x 3 matrix of alpha-GC connectivity for each of the two resting states.

### Statistical Analyses

Source-level power and GC were submitted to planned simple contrasts of States (S-RS vs. M-RS) to demonstrate the extent of alpha adaption from S-RS to M-RS. We then performed simple contrasts of Group (PTSD vs. Control) for the two states to test PTSD-related deficits in alpha power and GC, and double contrasts of State and Group to assess the effect of visual stimulation (S-RS minus M-RS) between groups. Pearson correlations were performed to assess clinical associations of alpha power and GC with symptom severity of hypervigilance. Guided by our previous surface-level analysis showing no difference between the two control (GAD and HC) (52), we combined them into a single Control group. In Supplemental Results, we confirmed that these two control groups did not differ in the source-level analyses.

Multiple-comparison corrections were applied for the analyses. For power analyses involving whole-brain voxel-wise comparisons, we used Monte Carlo simulations (with actual Gaussian filter widths extracted from the data) to derive the corrected threshold (*p* < .05): voxel level *p* < 0.005 (one-tailed) over eleven contiguous voxels. As for connectivity analyses, the 3 x 3 connectivity matrix resulted in 6 comparisons for each hypothesis testing, for which we applied the false discovery rate (FDR) criterion (FDR *p* < .05). Lastly, for clinical association analyses, we conducted confirmatory correlation analyses constrained to regions demonstrating main effects, for which correction was not applied. We also applied a whole-brain regression of alpha power on hypervigilance scores, followed by Monte Carlo multiple comparison correction. Trend-level and non-corrected effects will be reported but not further discussed.

## RESULTS

### PTSD-related alpha power deficits in the DMN and visual cortex

Validating intracranial source estimation of alpha oscillations, we observed “alpha blocking” by visual stimulation. Specifically, the contrast between S-RS versus M-RS (collapsed across groups) isolated alpha power reduction at M-RS across a large cluster (624 voxels) spanning bilateral visual cortices (peak voxel: −20, −100, 0; *t* = −4.50; Supplemental Figure 1A).

**Figure 1.**
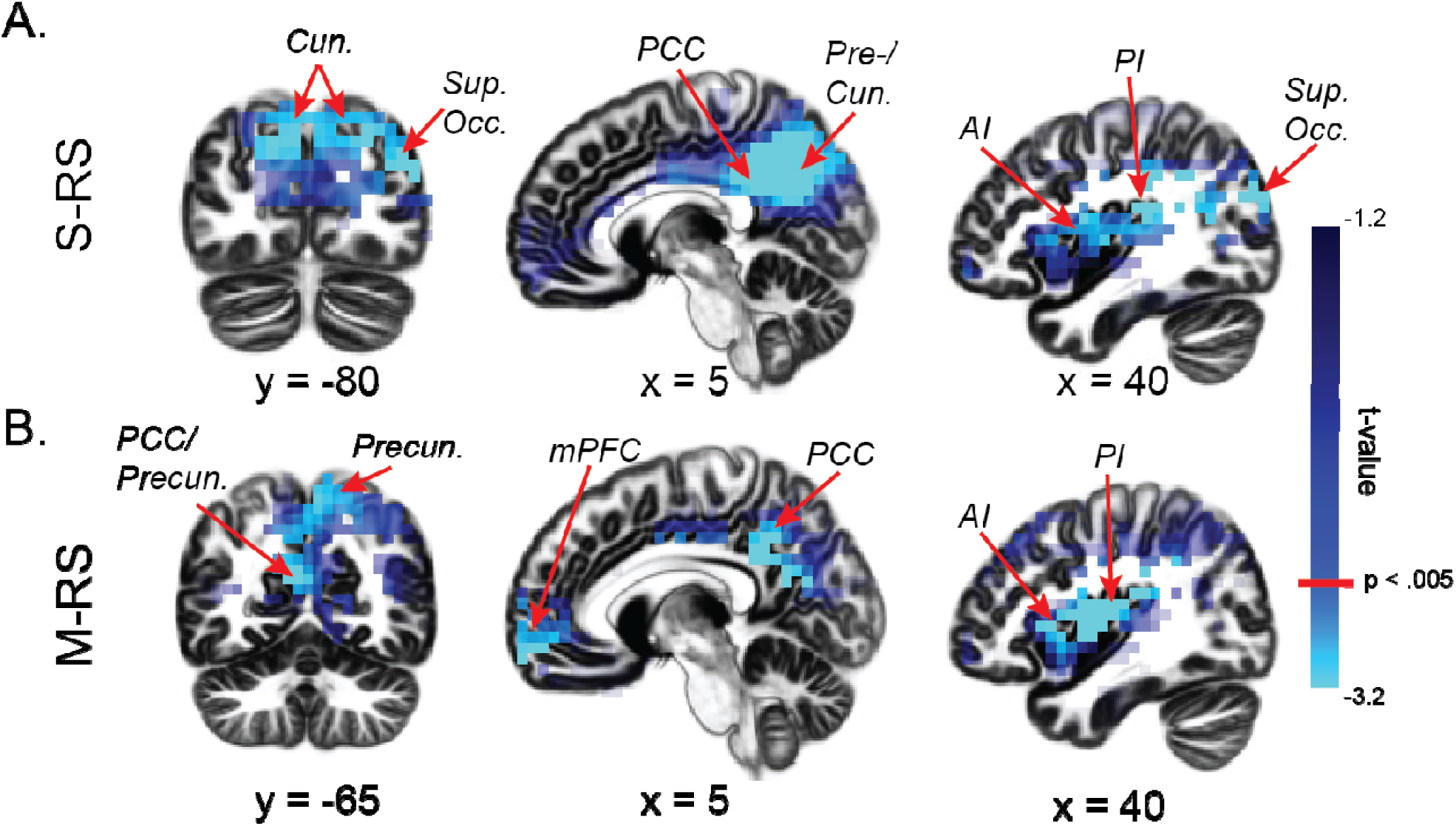
Group differences in alpha power. (A) During the S-RS, the PTSD group demonstrated reduced alpha power in the visual cortex (the cuneus, precuneus, and superior occipital gyrus), the posterior DMN hub (PCC), and anterior and posterior insula. (B) During the M-RS, the PTSD group showed reduced alpha power in the visual cortex (the cuneus and precuneus), both the anterior and posterior DMN hubs (mPFC and PCC), and anterior and posterior insula. Cun = cuneus; Precun = precuneus; AI = anterior insula; PI = posterior insula; Sup. Occ. = superior occipital gyrus.

Next, simple contrasts of Group (PTSD vs. Controls) revealed alpha power deficits in PTSD across the visual and DMN ROIs at each state. The visual cortex exhibited the strongest group effects: at S-RS, a large cluster (283 voxels) spanning bilateral cuneus (peaks: 5, −70, 30/-5, −70, 30; *t*’s < −3.41), bilateral precuneus (peaks: 5, −60, 35/-5, −65, 25; *t*’s < −3.75), and the right superior occipital gyrus (peak: 40, −80, 25; *t* = −3.90; Figure 1A); at M-RS, a cluster (68 voxels) in the bilateral precuneus, overlapping the S-RS cluster (peaks: 5, −55, 50/-5, −55, 40; *t*’s < −3.11; Figure 1B). Regarding the DMN, alpha deficits emerged in PTSD in the bilateral PCC for both states (S-RS: 60 voxels; peaks: 15, −45, 40/-5, −50, 35; *t*’s < −3.32; M-RS: 199 voxels; peaks: 5, −30, 25/-5, −30, 25; *t*’s < −3.76; Figure 1A-B), and the bilateral mPFC at M-RS only (88 voxels; peaks: 15, 60, −10/-15, 65, −15; *t*’s < −3.10; Figure 1B). Additional alpha power deficits (whole-brain corrected) appeared in the bilateral insula (S-RS: right/left: 49/76 voxels; peaks: 35, −5, 20/-35, 20, 5; Figure 1A; M-RS: right/left: 136/25 voxels; peaks: 40, −5, 15/-30, 25, 5; Figure 1B; *t*’s < −3.52). No significant clusters emerged for enhanced alpha power in the PTSD group, even at a lenient threshold of *p* < .05. Finally, double contrasts of State (M-RS minus S-RS) and Group (PTSD minus Control) yielded no difference between groups (voxel-level *p*’s > 0.11). These group effects are summarized in Table 2.

**Table 2.**
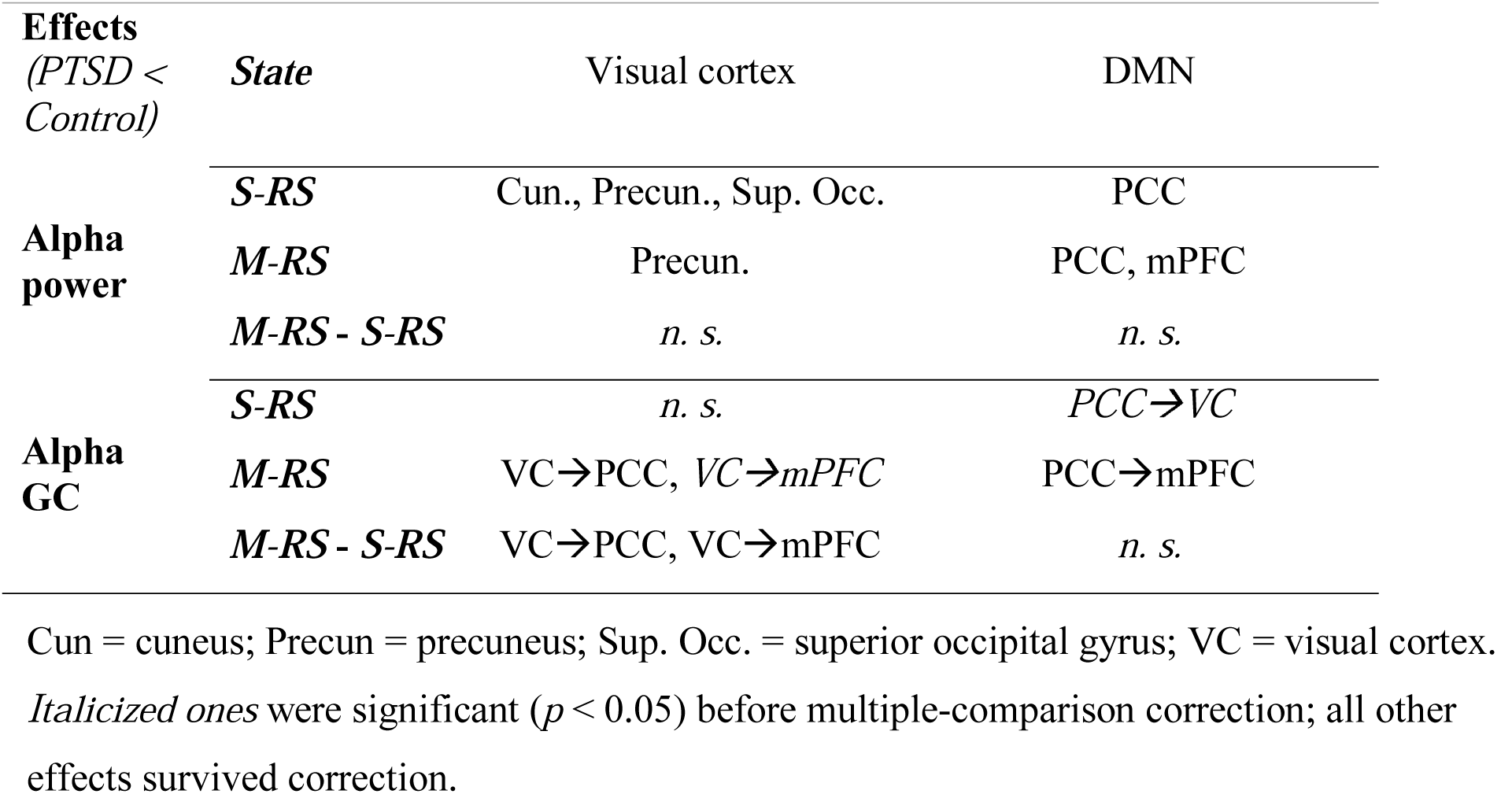
Summary of Group Effects.

### PTSD-related alpha connectivity deficits within and between the DMN and visual cortex

Like the power analyses above, we first assessed the effect of State (S-RS vs. M-RS) on alpha-frequency connectivity (collapsed across the groups). We observed reduced bidirectional alpha connectivity from the S-RS to the M-RS within the DMN: PCC →mPFC (M-RS minus S-RS; *t* = −3.49, *p* = 0.001, FDR *p* < 0.05) and mPFC→PCC (*t* = −3.14, *p* = 0.003, FDR *p* < 0.05; Supplemental Figure 1B), suggesting disrupted DMN connectivity in general by salient visual input (at M-RS).

Next, simple contrasts of Group for the S-RS revealed reduced alpha connectivity from the PCC to the visual cortex/VC (PCC →VC) in PTSD (vs. Controls; *t* = −2.26, *p* = 0.027; Figure 2A), albeit failing FDR correction. No other effects emerged during this state (*p*’s > .34). For the M-RS, the PTSD group (vs. Controls) demonstrated reduced connectivity within the DMN, including PCC →mPFC (*t* = −2.82, *p* = .008, FDR *p* < 0.05; Figure 2B) and, at a trend level, mPFC→PCC (*t* = −1.91, *p* = 0.064). The PTSD group demonstrated additional deficits in alpha connectivity from the visual cortex to both DMN ROIs at M-RS: VC→PCC (*t* = −3.06, *p* = 0.004, FDR *p* < 0.05) and VC→mPFC (*t* = −2.05, *p* = .049; albeit not FDR corrected). No effect appeared in the opposite (DMN→VC) direction (*p*’s > 0.12).

**Figure 2.**
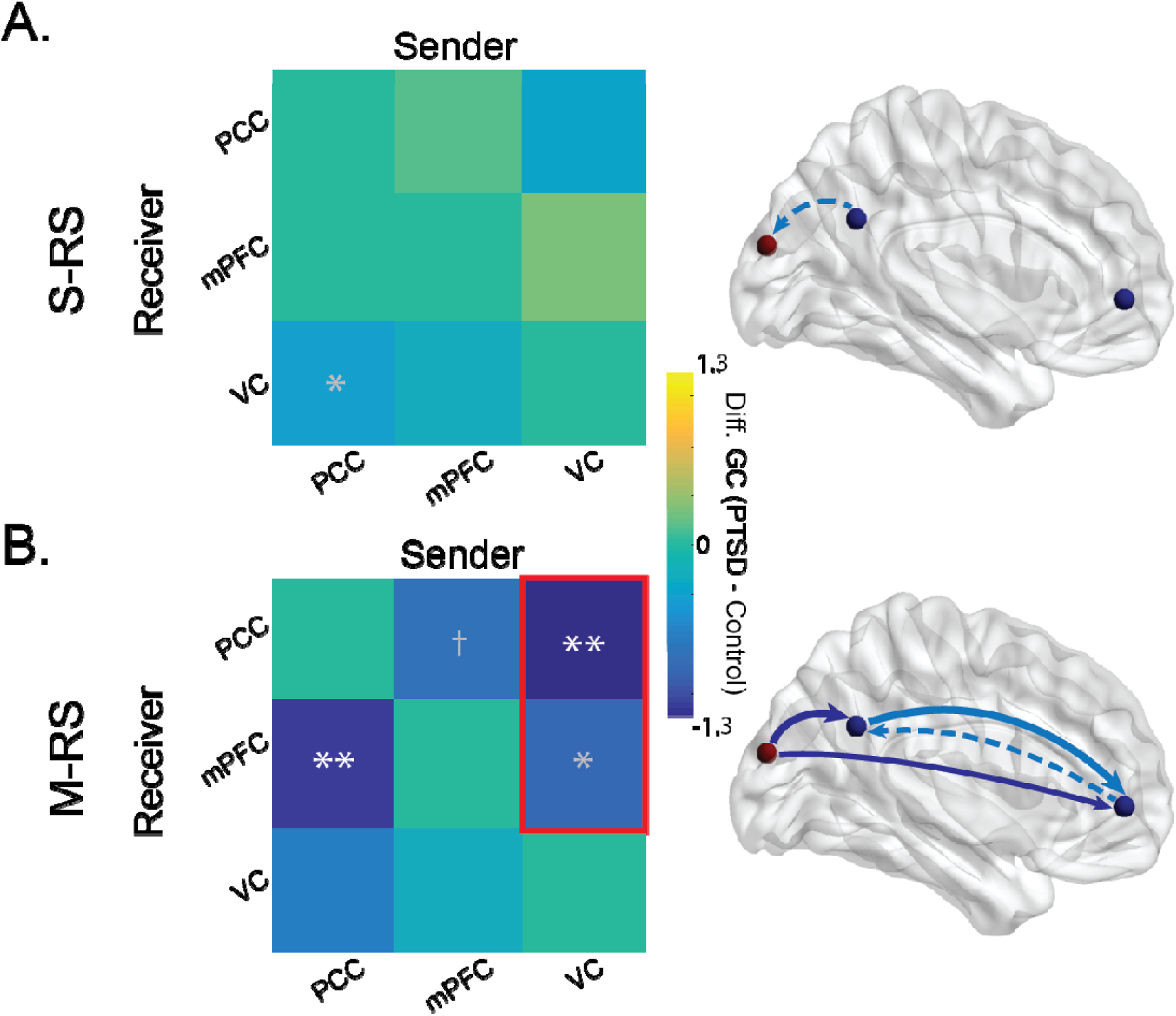
Group differences in alpha connectivity. Left column: matrices of group differences PTSD minus Controls) in directed alpha-frequency connectivity (GC) showed (A) reduced PCC→VC alpha connectivity (albeit not FDR corrected) during the S-RS; and (B) reduced PCC→mPFC alpha connectivity during the M-RS and, as enclosed in a red box, more reduction from the S-RS to the M-RS in VC→PCC and VC→mPFC alpha connectivity. Right column: Schematic presentations of group-differences in connectivity during the S-RS (A) and M-RS (B), with solid and dotted arrows reflecting connections surviving and not surviving FDR correction, respectively. Arrows in light blue and dark blue reflect significant effects from simple group contrasts and double contrasts of State and Group, respectively. Our discussion focused on the effects surviving the multiple comparison correction. \**p* < 0.05; \*\**p* < 0.01; † *p* < 0.1; white * = FDR corrected; gray * = not FDR corrected. VC = visual cortex.

Double contrasts of State (M-RS minus S-RS) and Group (PTSD vs. Control) revealed different effects of state (i.e., visual stimulation) on connectivity between the groups. Greater connectivity reduction from S-RS to M-RS appeared in the PTSD (vs. Control) group in VC→PCC (*t* = −2.22, *p* = 0.032) and VC→mPFC (*t* = −2.18, *p* = 0.036). Specifically, from S-RS to M-RS, significant connectivity reduction emerged in VC→PCC (*t* = −2.84, *p* = 0.011, FDR *p* < 0.05) and VC→mPFC (*t* = −3.04, *p* = 0.007, FDR *p* < 0.05) in the PTSD group, but none in the Control group (*p*’s > 0.628). These group effects are also summarized in Table 2.

### Clinical Associations

We then performed correlation analyses, regressing power or connectivity values on symptom severity in hypervigilance. Confirmatory analyses (constrained to the regions identified in the main contrasts) revealed negative associations between hypervigilance and alpha power in the precuneus/superior parietal lobule at S-RS (39 voxels; peak: −20, −75, −55; *r* = −0.49, *p* < .005; Figure 3A) and the DMN at M-RS: PCC (13 voxels; peak: 5, −45, 45; *r* = −0.28, *p* < .05) and mPFC (50 voxels; peak: −10, 65, −15; *r* = −0.30, *p* < .05; Figure 3B). Whole-brain regression analysis further revealed negative associations with alpha power in the ventral visual cortex at S-RS (left inferior temporal gyrus: 13 voxels; peak: −55, −5, −40; *r* = −0.40, *p* < .005 whole-brain corrected; Figure 3A). There was no significant correlation between hypervigilance and alpha connectivity (*p*’s > 0.20).

**Figure 3.**
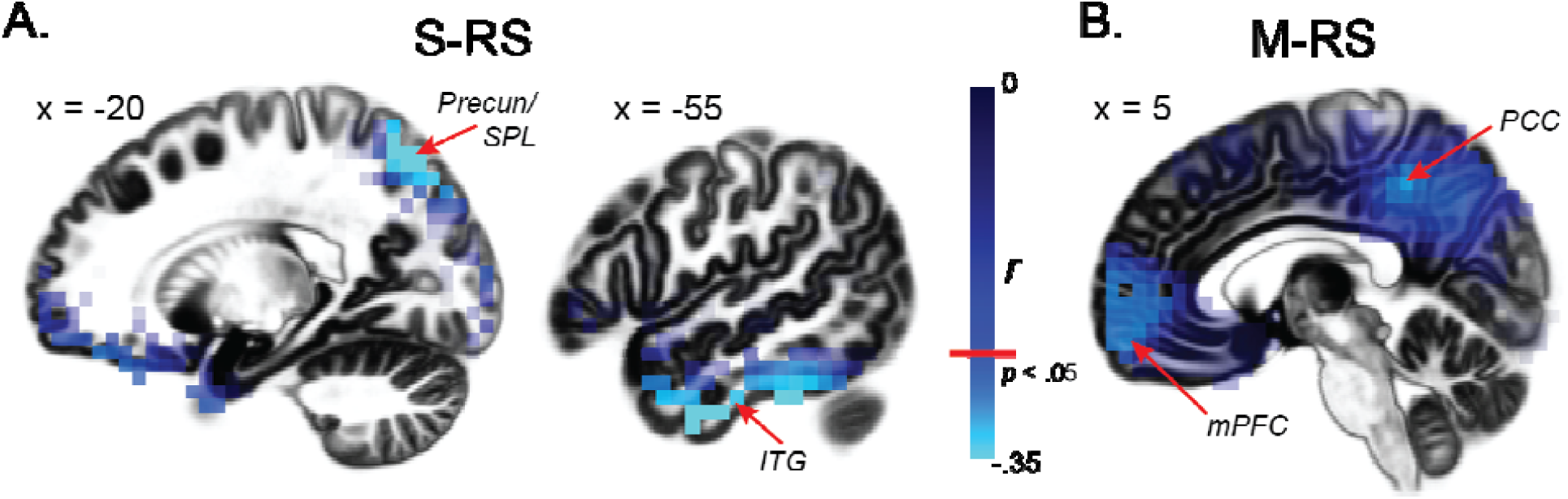
Clinical associations between alpha power and hypervigilance. Whole-brain correlation maps of alpha power and hypervigilance indicated negative correlations in both the dorsal (i.e., SPL, Precuneus) and ventral (i.e., ITG) visual cortices during the S-RS (A) and both DMN hubs (mPFC and PCC) during the M-RS (B). SPL = superior parietal lobule; ITG = inferior temporal gyrus.

## DISCUSSION

Source-level analysis of RS alpha oscillations isolated alpha (power and connectivity) deficits in the DMN and visual cortex in PTSD, especially during strong visual stimulation. In support of our first hypothesis — alpha deficits in the DMN, the PTSD group demonstrated reduced alpha power in the posterior DMN hub (the PCC) at both S-RS and M-RS as well as the anterior hub (the mPFC) at M-RS, accompanied by reduced DMN (PCC→mPFC) alpha-frequency connectivity at M-RS. In support of the second hypothesis — alpha deficits in the visual cortex, the PTSD group exhibited reduced alpha power in the visual cortex at both states. Importantly, joining alpha deficits in the two neural systems, diminished alpha-frequency connectivity from the visual cortex to the DMN was observed in the PTSD group at M-RS. Finally, alpha power deficits in the DMN and visual cortex directly correlated with symptom severity of hypervigilance. Therefore, linking anomalies in DMN and alpha activity in PTSD, the current results indicate interrelated alpha deficits in the DNM and visual cortex, implicating visual-cortex-DMN alpha dysrhythmia in the neuropathology of PTSD.

Neural oscillations actively participate in mental activities by modulating local neuronal excitability and mediating long-range neural communication (51, 74). Aberrant resting-state (intrinsic) neural oscillations (e.g., thalamocortical dysrhythmia) in neuropsychiatric disorders has been increasingly recognized (48, 49, 75), promoting the transdiagnostic conceptualization of “oscillopathies” for these disorders (50, 51). Advancements in neural computational algorithms (such as eLORETA) have permitted intracranial source estimation of neural oscillations in hdEEG recordings, providing important insights into oscillatory dysrhythmia in multiple disorders, e.g., schizophrenia (76, 77), depression (71), and PTSD (78). As validation of our hdEEG source analysis, we confirmed a strong “alpha blocking” effect of visual stimulation by demonstrating extensive alpha power reduction in bilateral visual cortices from the S-RS (minimal visual input) to the M-RS (strong visual input). By contrast, no other regions emerged from this contrast, highlighting the sensitivity and specificity of this source analysis of alpha oscillations.

Our source-level group analysis further identified alpha deficits within and between the visual cortex and the DMN in PTSD. Concerning the visual cortex, alpha oscillations are known to mediate visual cortical inhibition, and accordingly, alpha power correlates inversely with visual cortical activity (27, 28, 40, 41). In the PTSD group, reduced alpha power in the visual cortex was extensive and enduring across states. Without visual stimulation (at S-RS, reflecting intrinsic activity), alpha deficits spanned a large cluster over the cuneus, precuneus, and superior occipital gyrus, suggesting wide-spread, intrinsic neural disinhibition and hyperactivity across primary and secondary visual cortices in PTSD. This sensory cortical disinhibition aligns with extant electrophysiological evidence of impaired sensory gating and sensory cortical hyperactivity (to simple, neutral stimuli) (79-82) and behavioral disturbances in sensory filtering/gating and response in these patients (79, 83). With strong visual input at M-RS, alpha power reduction was particularly localized to the parietal visual cortex (primarily bilateral precuneus). Given that this region is strongly involved in visual spatial attention and perception (84-86) and that the M-RS condition mimics a real-life environment with salient sensory information, this alpha deficit (reflective of disinhibited visual spatial attention) can underlie hypervigilance in PTSD, expressed as excessive alertness to and scanning of the environment (87).

As mentioned above, alpha oscillations also play a critical role in long-range neural communication and is especially involved in the DMN; accordingly, alpha power correlates positively with DMN activity (32-39). The alpha deficit in the DMN hubs thus align with the extant literature citing DMN hypoactivity in PTSD (4, 7). The fact that DMN alpha power reduction extended from the PCC only at S-RS to both the PCC and mPFC at M-RS accentuates the particular DMN vulnerability in PTSD in a sensory-rich environment. In addition, as alpha oscillations synchronize activity and facilitate coherence across regions, reduced alpha power in these key DMN hubs further suggests compromised communication across the network in PTSD. Indeed, connectivity analyses revealed reduced PCC→mPFC alpha connectivity at M-RS in the PTSD group. This hypo-connectivity between the DMN hubs highlights impaired communication within the core architecture of the DMN. Clinically, DMN alpha deficits in both local power and inter-hub connectivity, especially acute in a sensory-rich environment, could contribute to difficulty in maintaining “tranquility” or “rest” (7, 17, 25) and avoidance of sensory stimulation in patients with PTSD (79, 83).

Our manipulation of visual stimulation between the two states revealed a direct link between alpha deficits in the visual cortex and the DMN. Consistent with general DMN susceptibility to salient sensory input, we confirmed a general reduction (in the entire sample) in bidirectional alpha connectivity between the DMN hubs (PCC→mPFC and mPFC→PC) as visual input increased from the S-RS to the M-RS. Beyond that, in the PTSD (but not Control) group, increased visual stimulation (from S-RS to M-RS) further diminished visual cortical alpha connectivity to both DMN hubs (VC→PCC and VC→mPFC). As mentioned above, alpha oscillations in the sensory cortex mediate sensory inhibition, such that this VC→DMN alpha projection would serve to protect the DMN by blocking sensory afferents to the network.

However, while this protective inhibitory process withstood the increased visual input at M-RS in the Control group, it broke down among patients with PTSD, suggesting impaired gating of sensory entry to (i.e., compromised protection of) the DMN. Exploratory correlation analyses further indicated a close correlation between VC→PCC and PCC→mPFC alpha connectivity at M-RS (*r* = 0.31, *p* = 0.013), highlighting a mechanistic link between these two pathways. That is, disinhibited visual cortical propagation to the PCC, in the presence of strong environmental input, could further impair PCC-driven alpha synchronization with the mPFC, worsening DMN dysfunction in PTSD.

Together, current findings implicate a core visual-cortex-DMN system of alpha dysrhythmia in PTSD. The critical role of visual cortical alpha deficits in this pathology lends credence to our sensory hypothesis of PTSD centered on sensory cortical disinhibition (52, 88) and bottom-up accounts of PTSD in general (89, 90). This core visual-cortex-DMN system of alpha dysrhythmia was reinforced by the specificity of alpha deficit sources. Despite the use of whole-brain analyses, group differences in alpha power and clinical associations between alpha power and hypervigilance were localized to the visual cortex and the DMN only (except for the insula as discussed below). While DMN hypoactivity is known to be associated with PTSD symptom severity (5, 7, 14, 91), the clinical association in the visual cortex highlights an additional pathway linking visual cortical disinhibition to PTSD symptoms. Notably, the associations with hypervigilance implicate not only both the dorsal (i.e., the parietal cortex) and ventral visual cortices (i.e., the inferior temporal gyrus) but also the higher-order regions with strong interactions with limbic and frontal regions. As the ventral visual cortex underpins visual object perception, its involvement here aligns with the fact that hypervigilance in PTSD associated with hyperactivity and hypersensitivity to (threat and neutral) sensory cues (92, 93), in addition to exessive spatial scanning engaging the doral visual cortex.

While the insula was not an *a priori* ROI in the current study, expansive alpha deficits spanning the posterior and anterior insula emerged in PTSD following whole-brain correction. The insula is a highly heterogeneous structure, with the posterior portion receiving strong sensory afferents and the anterior portion densely connected with limbic and prefrontal regions as a key node of the salience network/SN (94-97). Abnormal insular and SN activity has been repeatedly observed in patients with PTSD (4, 15), and alpha suppression via neurofeedback can increase SN activity (98). Our data provide preliminary evidence of insular alpha dysrhythmia in PTSD, potentially underlying some of the insular anomalies.

We included patients with GAD as an additional control condition to rule out general effects of anxiety and arousal on alpha activity. Our previous sensor-level analysis of alpha oscillations revealed no alpha deficits in this group (52), which directed us to combine patients with GAD and healthy controls into one control group in the source-level analysis. Nonetheless, as reported in Supplemental Results, we systematically examined source-level anomalies in the GAD group, confirming no difference between them and healthy controls, except for enhanced mPFC→PCC alpha connectivity at M-RS. We surmise that this mPFC→PCC hyper-connectivity could heighten DMN functioning in GAD, supporting the hallmark symptoms of self-referential rumination and worry in these patients. Overall, these results ruled out the possible confound of general anxious arousal in attenuating alpha activity, highlighting a specific alpha dysrhythmia in PTSD.

Integrative neuroimaging and electrophysiological research have promoted the idea that alpha oscillations sustain and facilitate DMN functioning by synchronizing spontaneous activity across the network and suppressing sensory cortical propagation. Translating these mechanisms to the neuropathology of PTSD, we confirmed interconnected alpha deficits in the DMN and visual cortex in patients with PTSD. Therefore, the current findings provide the first evidence of visual-cortex-DMN alpha dysrhythmia in PTSD, presenting a unifying neural underpinning of sensory disinhibition, DMN dysfunction, and hypervigilance in this disorder. The specification of this alpha dysrhythmia further isolates a novel therapeutic target, promoting network-based interventions (15) using brain stimulation of alpha oscillations (99) in the visual-cortex-DMN system as a new line of treatment for PTSD.

## Data Availability

Data may be made available upon request.

## Acknowledgement

This research was supported by the National Institute of Mental Health grants R01MH093413 (W.L.), the FSU Chemical Senses Training (CTP) Grant Award T32DC000044 (K.C.) from the National Institutes of Health (NIH/NIDCD), and the Subaward of U.S. Army award W81XWH-10-2-018 (N.S.), which does not necessarily represent the views of the Department of Defense, Department of Veterans Affairs, or the United States Government, nor does it constitute or imply endorsement, sponsorship, or favoring of the study design, analysis, or recommendations..

## Competing financial interests

The authors declare no competing financial interests or potential conflicts of interest.

## SUPPLEMENTARY METHODS

### FASTER Algorithm

The *Fully Automated Statistical Thresholding for EEG artifact Rejection* (FASTER) algorithm was used for the detection and removal of artifacts within the data. Using a z-score threshold of ± 3, the FASTER algorithm detects and corrects for artifacts within single channels, individual epochs, independent components, and within-epoch channels. FASTER first interpolates deviant channels from the continuous data using the EEGLAB spherical spline interpolation function. Data were then segmented into non-overlapping, 1-second long, mean-centered epochs. Epochs were then rejected based on z-scores ± 3 within parameters of amplitude range, variance, and deviation. Epoched data were then re-referenced to averaged mastoids and submitted to independent component analysis (ICA) decomposition using the Infomax algorithm. Artefactual components (i.e. muscular artifacts, eye blinks and saccades, electrode “pop-offs”, etc.) were automatically detected and removed from the data. Lastly, deviant channels within individual, cleaned epochs were interpolated again using the EEGLAB spherical spline interpolation function.

## SUPPLEMENTARY RESULTS

### Group contrasts of alpha power

Alpha power contrasts of PTSD versus healthy controls (HC) and GAD controls, individually, revealed consistent results to those reported in the main text (Supplemental Figure 2). At S-RS, relative to HC, patients with PTSD demonstrated reduced alpha in a large (71 voxel) cluster spanning the PCC/precuneus (peak: 20, −60, 35) and cuneus/superior occipital gyrus (peak: 45, −80, 25; *t*’s < −3.42). A contrast of PTSD vs. GAD revealed a similar cluster (112 voxel) spanning the PCC/precuneus (peak: 15, −50, 40) and cuneus (peak: 5, −65, 15; *t*’s < 3.31). Finally, patients with PTSD demonstrated reduced alpha in the right insula compared to HC (10 voxels; peak: 35, −5, 15; *t*’s < −3.18) and left insula compared to GAD (30 voxels; peak: − 30, 25, 0; *t*’s < 3.29).

At M-RS, patients with PTSD demonstrated similar reductions in alpha power over a large cluster spanning the PCC/precuneus, relative to HC (121 voxels; peak: 5, −55, 50; *t*’s < − 2.41) and GAD (112 voxels; peak: 15, −45, 40; *t*’s < −3.31). An additional cluster emerged in the mPFC, relative to HC (10 voxels; peak: 10, 65, 0; *t*’s < 2.87) and GAD (95 voxels; peak: −15, 65, −15; *t*’s < −3.31), consistent with results reported in the main text spanning the posterior and anterior hubs of the DMN. Additional deficits emerged in the right insula compared to HC (10 voxels; peak: 45, −5, 15; *t’s* < −2.98) and bilateral insula compared to GAD (right/left: 84/65 voxels; peak: 60, −15, 30/-30, 25, 5; *t*’s < −3.58). Notably, there were no significant differences between HC and GAD groups at either state.

### Group contrasts of alpha-frequency connectivity

Alpha connectivity contrasts of PTSD versus healthy controls (HC) and GAD controls, individually, revealed consistent results to those reported in the main text (Supplemental Table 1). At S-RS, a marginal decrease in alpha connectivity from PCC →VC emerged in PTSD (vs. HC: *t* = −1.72, *p* = 0.092; vs. GAD: *t* = −1.83, *p* = 0.073). At M-RS, PTSD similarly showed decreased alpha connectivity from PCC →mPFC (vs. HC: *t* =-2.78, *p* = 0.008; vs. GAD: *t* = −2.68, *p* = 0.010), as well as from VC → PCC (vs. HC: *t* = −2.36, *p* = 0.023; vs. GAD: *t* = −3.30, *p* = 0.002). Notably, HC and GAD did not differ on any of these contrasts (*p*’s > 0.224). However, relative to both PTSD and HC, the GAD group demonstrated increased alpha connectivity from mPFC → PCC (PTSD vs. GAD: *t* = 3.25, *p* = 0.003; HC vs. GAD: *t* = 2.65, *p* = 0.012), suggesting GAD-specific augmentation in alpha connectivity.

Double contrasts of State (M-RS minus S-RS) and individual Group additionally revealed marginally greater decreases in VC→PCC alpha connectivity from the S-RS to M-RS in PTSD relative to HC (*t* = 1.87, *p* = 0.068) and GAD (*t* = −1.87, *p* = 0.069). Furthermore, greater decreases in VC→mPFC alpha connectivity emerged from the PTSD versus GAD contrast (*t* = − 2.29, *p* = 0.027), but not for the PTSD versus HC contrast (*t* = −1.50, *p* = 0.142), albeit in the right direction. HC and GAD did not differ on any of these contrasts (*p*’s > 0.411).

**Supplemental Table 1.** Participant Demographics.

|  | **PTSD** | **GAD** | **HC** |
| --- | --- | --- | --- |
| *n* | 25 | 24 | 20 |
| Age (years) | 34.6 ± 10.4 | 30.0 ± 12.5 | 32.5 ± 13.8 |
| Gender (female/male) | 16/9 | 17/7 | 8/12 |
| Substance Use (%) † | 64%*** | 0 | 15% |
| Medication Use (%) | 40% | 50% | 20% |
| PCL Total | 61.0 ± 16.1** | 43.2 ± 14.0** | 30.7 ± 9.0 |
| PCL - Hypervigilance | 4.1 ± 1.0** | 2.4 ± 1.3* | 1.7 ± 0.8 |
| BAI | 26.2 ± 15.7* | 15.8 ± 9.1* | 7.0 ± 7.5 |
| BDI | 26.8 ± 12.5** | 22.3 ± 7.5** | 11.6 ± 8.8 |
| **Axis I Comorbidity** | *n* (%) | *n* (%) | — |
| No comorbid diagnoses | 4 (16%) | 10 (42%) | — |
| Comorbid diagnoses | 21 (84%) | 14 (58%) | — |
| PDD | 12 (48%) | 3 (13%) | — |
| SAD | 9 (36%) | 8 (33%) | — |
| MDD | 5 (20%) | 3 (13%) | — |
| Panic Disorder | 3 (12%) | 1 (4%) | — |
| Specific Phobia | 2 (8%) | 3 (13%) | — |
| OCD | 2 (8%) | 0 | — |
| Alcohol Use | 3 (12%) | 0 | — |
| Substance Use | 3 (12%) | 0 | — |
| Other‡ | 10 (40%) | 5 (21%) | — |
| Mean # Dxns (SD) | 2.22 (1.47) | 1.00 (1.25) | — |

BAI = Beck Anxiety Inventory; BDI = Beck Depression Inventory; PCL = Posttraumatic Stress Disorder Checklist; PDD = Persistent Depressive Disorder; SAD = Social Anxiety Disorder; OCD = Obsessive Compulsive Disorder; † = Subjects with opioid, stimulant and cocaine use were excluded. \* $p < .05$ , \*\* $p < .01$ , \*\*\* $p < .005$ ;
‡ Other diagnoses refer to diagnoses endorsed by no more than 1 subject per group, including: Binge Eating Disorder, Agoraphobia, Hoarding, Depressive-Not Otherwise Specified, Bulimia, Trichotillomania, Excoriation, Anxiety-Not Otherwise Specified, Anorexia Nervosa, and Other Trauma-Related Disorders.

**Supplemental Table 2.** Group contrasts of alpha-frequency connectivity.

|  | PTSD vs. HC | PTSD vs. GAD | HC vs. GAD |
| --- | --- | --- | --- |
| **S-RS:** |  |  |  |
| PCC → VC | $t = -1.72, p = 0.092$ | $t = -1.83, p = 0.073$ | *n.s.* |
| **M-RS:** |  |  |  |
| PCC → mPFC | $t = -2.78, p = 0.008$ | $t = -2.68, p = 0.010$ | *n.s.* |
| VC → PCC | $t = -2.36, p = 0.023$ | $t = -3.30, p = 0.002$ | *n.s.* |
| mPFC → PCC | *n.s.* | $t = -3.25, p = 0.003$ | $t = -2.65, p = 0.012$ |
| **M-RS minus S-RS:** |  |  |  |
| VC → PCC | $t = -1.87, p = 0.068$ | $t = -1.87, p = 0.069$ | *n.s.* |
| VC → mPFC | *n.s.* | $t = -2.29, p = 0.027$ | *n.s.* |

## SUPPLEMENTARY FIGURES

**Supp. Figure 1.**
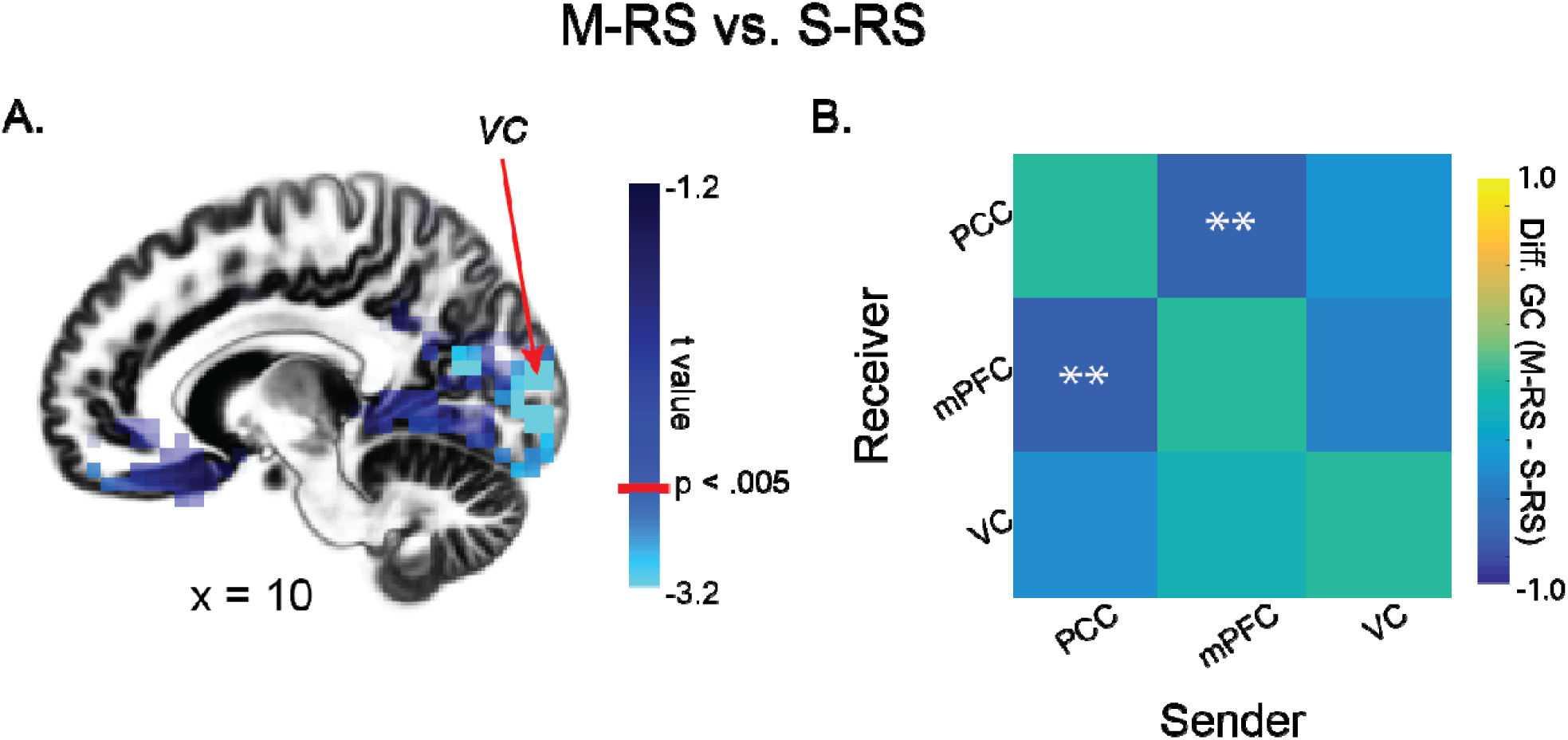
State effects: differences between states (M-RS minus S-RS collapsed across groups) in alpha power (A) and alpha-frequency GC connectivity (B). ** *p* < 0.01.

**Supp. Figure 2.**
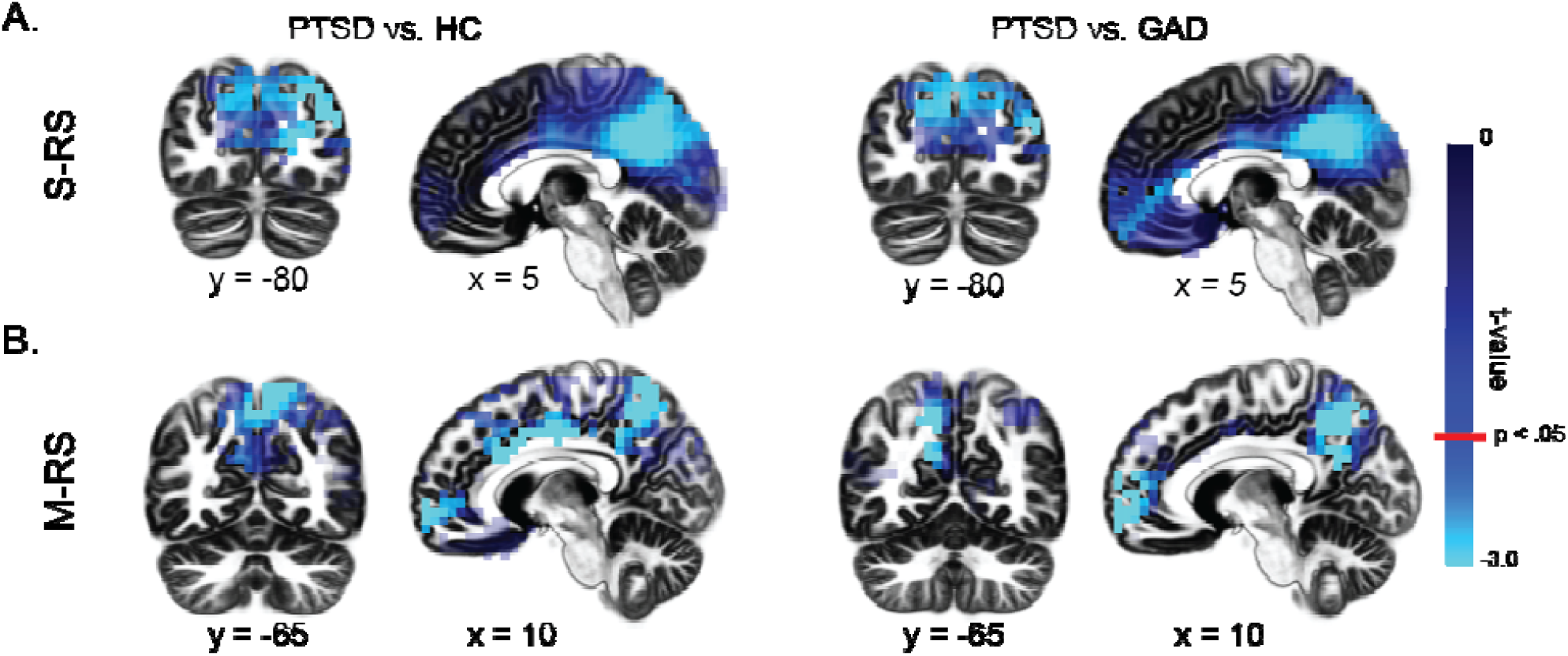
eLORETA contrasts of alpha power for PTSD vs. Healthy Controls and GAD, individually, for the S-RS (A) and M-RS (B).

## Notes

### Competing Interest Statement

The authors have declared no competing interest.

